# Mobile-linked point-of-care diagnostics in community-based healthcare: A scoping review of user experiences

**DOI:** 10.1101/2023.05.28.23290656

**Authors:** Siphesihle R. Nxele, Boitumelo Moetlhoa, Thobeka Dlangalala, Kuhlula Maluleke, Kabelo Kgarosi, Ashleigh B. Theberge, Tivani Mashamba-Thompson

**Author notes:** Corresponding author Siphesihle R. Nxele.

## Abstract

**Background:** While mobile-linked point-of-care diagnostics may circumvent geographical and temporal barriers to efficient communication, the use of such technology in community settings will depend on user experience. We conducted a scoping review to systematically map evidence on user experiences of mobile-linked point-of-care diagnostics in community healthcare settings.

**Methodology:** We conducted a comprehensive search of the following electronic databases: Scopus, Web of Science, and EBSCOhost (Medline, CINAHL, Africa-wide, Academic Search Complete). The inter-reviewer agreement was determined using Cohen’s kappa statistic. Data quality was appraised using the mixed method appraisal tool version 2018, and the results were reported according to the preferred reporting items for systematic reviews and meta-analyses for scoping reviews (PRISMA-ScR).

**Results:** Following the abstract and full article screening, nine articles were found eligible for inclusion in data extraction. Following the quality appraisal, one study scored 72.5%, one study scored 95%, and the remaining seven studies scored 100%. Inter-rater agreement was 83.54% (Kappa statistic = 0.51, p < 0.05). Three themes emerged from the articles: approaches to implementing mobile-linked point-of-care diagnostics, user engagement in community-based healthcare settings, and limited user experiences in mobile-linked point-of-care diagnostics. User experiences are key to the sustainable implementation of mobile-linked point-of-care diagnostics. User experiences have been evaluated in small community healthcare settings. There is limited evidence of research aimed at evaluating the usability of mobile-linked diagnostics at the community level.

**Conclusion:** More studies are needed to assess the user experience of mobile-linked diagnostics in larger communities. This scoping review revealed gaps that need to be addressed to improve user experiences of mobile-linked diagnostics, including language barriers, privacy issues, and clear instructions.

## Introduction

Diagnostics plays an important role in disease management and prevention [1]. Technological advancements have led to the development of mobile-linked (m-linked), point-of-care (POC) diagnostics, which have revolutionized access to medical care by enabling rapid testing, early detection, and diagnosis regardless of geographical location [2, 3]. Using mobile-linked POC diagnostics, medical personnel can process, transfer, and interpret data quicker, allowing for better decision-making [4]. In community healthcare settings, mobile-linked POC diagnostics can alleviate the burden on healthcare systems, as evidenced during the COVID-19 pandemic [5]. Effective diagnostics may reduce the number of hospitalizations and prevent premature mortality. The development and implementation of POC diagnostics for human immunodeficiency virus (HIV), tuberculosis (TB), and malaria screening have significantly reduced morbidity and mortality in developing settings where laboratory infrastructure is lacking [1].

The Joint United Nations Program on HIV/AIDS (UNAIDs) and the World Health Organization (WHO) have worked on strategies to link wireless mobile communications to POC diagnostics to ultimately improve healthcare systems [6]. The WHO also compiled an assessment to discuss upscaling of mHealth innovations for women, children, and adolescent health [7]. However, to upscale, the technological experiences, lifestyle, and general behaviors of end users in community settings would need to be understood [8].

The sustainability of m-linked POC diagnostics in community settings depends greatly on the end users [9]. The widespread use of smartphones has enabled the ability to collect data on user preferences, and these data can be incorporated into the development of m-linked POC diagnostic health interventions [10]. For a positive user experience, technology should be designed to meet the needs and requirements of users, who should be involved in the development process [11]. Currently, the evidence on user experiences with m-linked POC diagnostics in community settings is unclear. In this scoping review, we systematically map evidence on user experiences of m-linked POC diagnostics in community settings. This scoping review will guide future research on how users experience m-linked POC diagnostics.

## Methodology

### Study design

This scoping review was conducted in line with the methodological framework proposed by Arksey and O’Malley [12] and further advanced by Levac et al. [13]. We chose a scoping review as we wanted to investigate the extent of information available to ultimately identify knowledge gaps [14]. The protocol is registered on Open Science Framework (https://archive.org/details/nxele-a-scoping-review-protocol-on-integration-of-mobile-linked-poc-diagnostics).

### Identifying the research question

We used the PCC (Population, Concept, and Context) nomenclature to conceptualize the research question (**Table 1**). The research question for this scoping review is: What are the user experiences of m-linked POC diagnostics in community-based healthcare?

**Table 1.** PCC for determining the eligibility of the research question

|  |  |
| --- | --- |
| Population | m-linked POC diagnostics |
| Concept | User experience in health technologies |
| Context | Community based healthcare |

**Population**: m-linked POC diagnostics is defined as technology that allows for screening and diagnostics of communicable and non-communicable diseases in remote settings by healthcare professionals and patients [6] such as COVID-19 tests [15] and chest x-ray evaluations [16]. This technology would shorten the time between testing and clinical diagnosis [17].

**Concept**: user experience in health technologies refers to how users interact with the technology and their response to it.

**Context**: Community-based healthcare is defined as healthcare in targeted populations, which involves providing healthcare services on a local, personalized level [18]. An example of community-based healthcare would include community healthcare facilities that provide services such as community nursing, aged care, and occupational therapist services [19].

### Identifying relevant studies

We conducted a comprehensive and reproducible literature search of the following electronic databases: Scopus, Web of Science, and EBSCOhost (Medline, CINAHL, Africa-Wide, Academic Search Complete). The principal investigator (SRN), subject specialist (TM-T), and information specialist (KK) developed a comprehensive search strategy to ensure the correct use of indexing terminology and Medical Subject Headings [20] [20]. We snowball-searched the references cited in the included studies to identify studies not indexed in electronic databases. We did not apply any language restrictions to minimize the risk of excluding relevant studies. The following keywords were searched and refined to suit each database: 1, User experience, and user experience in health technologies; 2, mobile-linked point-of-care diagnostics; and 3, community-based healthcare. The search string comprises a set of keywords connected by Boolean operators, “AND,” “OR,” brackets, and quotations. Each database search was documented in detail, showing the keywords, date of search, electronic database, and the number of retrieved studies, and the results of the search were tabulated in a search summary table (**S1 Table).**

### Eligibility criteria

#### Inclusion criteria

We included articles reporting on:

- m-linked POC diagnostics in a community-based healthcare setting
- technology that allows for screening and diagnostics of communicable and non-communicable diseases in remote settings
- user experiences in health technologies
- evidence of community-based healthcare on a local, personalized level

#### Exclusion criteria

We excluded articles that:

- lacked evidence on POC diagnostics in community-based healthcare
- lacked evidence on the user experiences of m-linked POC diagnostics
- Articles pre-dating the year 2016
- Review articles

### Selection of eligible studies

All eligible articles were imported into Endnote X7. We removed duplicates before the title and abstract screening phase. The titles and abstracts were screened using Rayyan software and guided by the eligibility criteria. This was followed by full article screening which was aided by Google Forms and guided by the eligibility criteria. Discrepancies in abstract screening were resolved through a discussion with the project team. Discrepancies in full article screening were resolved by a third reviewer. Inter-reviewer agreement after full article screening was tabulated **(S2 Table)** and expressed using Cohen’s kappa coefficient (κ) statistic using Stata 13.0SE (StataCorp College Station, TX, USA) [21] **(S1 File)**. The kappa statistics results were interpreted as follows: values < 0.1 indicate no agreement, 0.10–0.20 indicate none to slight, 0.21–0.40 as fair, 0.41–0.60 as moderate, 0.61–0.80 as substantial, and 0.81–1.00 as almost perfect agreement.

### Charting data

We extracted data from the included studies using a piloted form designed in Google Forms. The following information was extracted from included studies: author(s) and date, title, aim, country, study/community-based healthcare setting, study population, m-linked POC diagnostic tool, user experience, and main findings.

### Ethical considerations

This scoping review synthesized the existing literature. Therefore, a review of the proposal by the ethics committee was not required.

### Quality appraisal

We evaluated the quality of the included articles using the mixed method appraisal tool [22] version 2018 [23]. Using MMAT, the methodological quality of five categories of research was appraised, which included the following: qualitative research, randomized controlled trials, nonrandomized studies, quantitative descriptive studies, and mixed methods studies (**S3 Table**). The quality of evidence was represented as follows: (i) ≤ 50%, low-quality evidence; (ii) 51–75%, average-quality evidence; and (iii) 76–100%, high-quality evidence.

### Summary and collating

The extracted data were thematically analyzed. The emerging themes were then summarized.

## Results

### Screening results

The initial search yielded 981 articles. After title and abstract screening, 79 studies remained (**Fig 1**). After the full-text screening, nine articles were deemed eligible for data extraction. The excluded studies and reasons for exclusion are given in **S4 Table.** The inter-rater agreement was high (83.54%, K = 0.51, p < 0.05). In addition, McNemar’s Chi-square statistic suggests that there was no significant difference in the proportions of yes/no answers by the reviewers (p > 0.01).

**Fig. 1:**
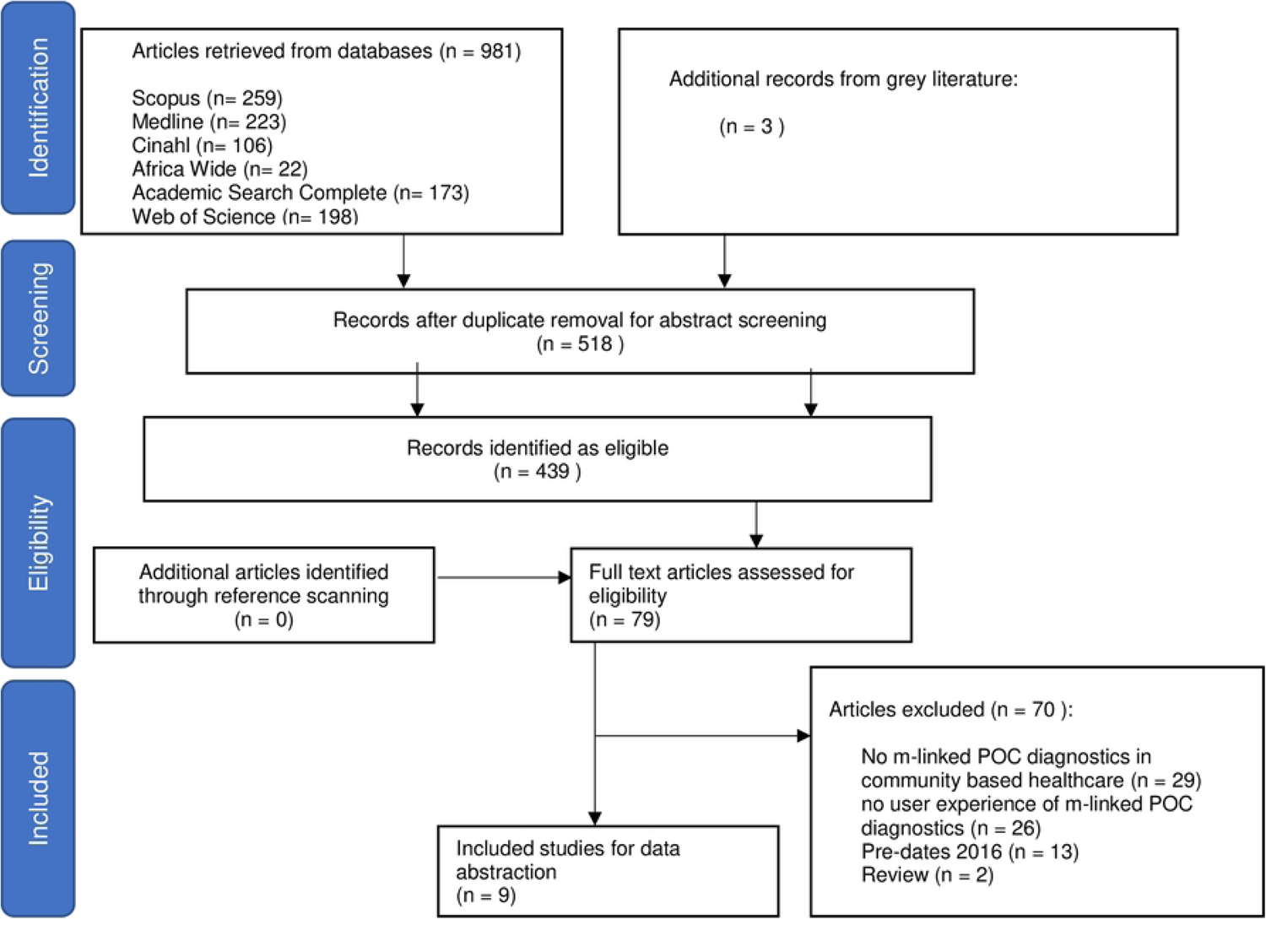
Preferred Reporting Items for Systematic Reviews and Meta-Analyses for scoping reviews flow diagram showing results of literature search and screening.

### Characteristics of the included studies

The characteristics of the included articles are detailed in **Table 2**. These studies were published between 2016 and 2022. The studies presented evidence on user experiences of m-linked POC diagnostics. The included articles comprised two cross-sectional studies [24, 25], two qualitative studies [26, 27], a cohort study [28], three surveys [29–31], and one mixed method study [32].

**Table 2:** Characteristics and findings of the studies included in the scoping review.

| Author(s)<br>(year) | Title | Aim | Country | Study design | m-linked POC diagnostic tool | Community setting and study population | User experience | Main findings | Reference |
| --- | --- | --- | --- | --- | --- | --- | --- | --- | --- |
| Asgary et al. (2019) | Acceptability and implementation challenges of smartphone-based training of community health nurses for visual inspection with acetic acid in Ghana: mHealth and cervical cancer screening. | To explore the acceptability and feasibility of smartphone-based training in cervical cancer screening using visual inspection with acetic acid (VIA)/cervicography. | Ghana | Qualitative | Smartphone POC diagnostic tool | Community health centers (CHCs) in Accra<br>Community health nurses (CHNs), patients, nurse supervisor, expert reviewer | CHNs agreed that learning pelvic anatomy was interesting. CHNs found smartphones easy to use for cervicography and sending photos to the mentor. Nurses did not face any major technical challenges in performing VIA. Few patients understood the concept of screening and prevention. Most patients appreciated cervical photography | Patients accepted smartphone-based VIA. Neither group cited significant barriers to performing or receiving VIA at CHCs. Could incorporate smartphone imaging and mentorship via text messaging. CHNs were able to leverage their existing community relationships to address a lack of knowledge and misperceptions. Findings limited to CHCs. | [26] |
| Farao et.al. (2020) | A user-centred design framework for mHealth | To explore a user-centered approach for mHealth design in a developing, under-resourced context. | South Africa | Cross sectional | m-linked POC tuberculin skin test (TST) | Developing and under-resourced setting<br>Graduate students | Participants struggled to follow and remember instructions. Unclear video instructions without audio assistance. Misunderstanding of continuous, scrollable app pages and disjointed ones. Difficulty capturing multiple images at different angles. Uncertainty when submitting images. | This study did not engage the intended end users throughout the study. Small sample size limited generalisability. The use of the PSSUQ in English was a limitation with a subgroup of users who preferred isiXhosa as their spoken language and language of instruction. | [24] |
| Jacobson et.al. (2020) | Feasibility of integrating a mobile decision-support app into a multicomponent CME initiative. Developing clinician competence at the point of care | To describe the development and incorporation of a mobile application into multiple activity formats within the European Immuno-Oncology Clinic Companion CME initiative. | Global 10 countries | Cohort | Mobile application on smartphone | European immuno-oncology clinic CME community<br>All ONCOassist users | No integration of electronic medical record (EMR), incomplete information provided | Poor engagement with some features of the Toxicity Tool. The share and favorite features were used 191 and 147 times, respectively. Clinicians accessed the info screen, which described the methods for irAE management algorithm development and linked to the EIOCC CME portal, 690 times. | [28] |
| Kadam et al. (2020) | Target product profile for a mobile app to read rapid diagnostic tests to strengthen infectious disease surveillance | To develop a Target Product Profile (TPP) for a mobile phone app for a rapid diagnostic test. The app is intended to transmit RDT test data, patient data, and contextual data. | UK | Survey | Mobile phone application | Infectious disease testing sites<br>Clinical users, healthcare programs, health information systems, surveillance systems, and global public health stakeholders. | Need for additional language support. Several RDTs should run at one time. Security and privacy: The EU's GDPR cannot be applied as a whole outside of the EU. | Stakeholders agreed that the app should respect local approaches to data ownership, security, and storage. The app to be free to use. | [29] |
| Kerwagen et al. (2022) | Point-of-care speech-recognition based mHealth solution to facilitate physicians' daily work. | To quantify the user experience of a point-of-care speech-recognition based mHealth solution | Germany | Survey | Speech-recognition based mHealth solution | Department of Trauma and Plastic Surgery at the University Hospital Würzburg<br>All physicians | User friendly | Participants supported the mHealth solution. The mHealth solution was perceived as supportive, helpful, easy to use, inventive, and leading edge. The POC diagnostic tool was acceptable to the user and user friendly. | [30] |
| Lavallee et al. (2019) | Engaging patients in co-design of mobile health tools for surgical site infection surveillance: Implications for research and implementation | To explore the perspectives of participants on benefits, value, and potential drawbacks of mHealth | USA | Qualitative | Surgical site infection surveillance | Post-operative recovery<br>Key stakeholders | Stakeholders suggested a need for increased patient involvement to improve POC diagnostics | In the context of post-operative surgical care and SSI surveillance, patients and healthcare stakeholders should be involved in the co-design of new technology. Research funding should align with the needs of healthcare decision-makers. | [27] |
| Redley et al. (2019) | Co-development of "BRAIN-TRK": Qualitative examination of acceptability, usability, and feasibility of an App to support nurses' care for patients with behavioural and psychological symptoms of neurocognitive disorders in hospital settings | (a) Describe the co-development of a point-of-care app (b) Report the acceptability, usability, and feasibility of the app | Australia | Mixed-methods | App to support nurses' care for patients with behavioural and psychological symptoms of neurocognitive disorders | Hospital settings<br>Nurses | The app was acceptable. Nurses were familiar with the app and recognized the benefits. Nurses felt that it would increase workload. Other barriers included inconsistent use, pressure to use, and resistance to change. | Nurses used the app with 44.7% (n = 17) of eligible inpatients. Cognitive screening was completed at least once for each patient, with 146 risk assessments recorded. Acceptability was enhanced by easy navigation, and clear and useful content, but hindered by unclear expectations, unfamiliarity, and device-related factors. | [32] |
| Shin et al. (2019) | Healthcare worker feedback on a prototype smartphone-based point-of-care test platform for use in episodic care | To evaluate the end-user acceptability of an experimental POC test platform with novel technical features. | USA | Cross-sectional survey | Mobile nucleic acid amplification tests (NAAT) for testing chlamydia | Urban academic emergency department<br>Socioeconomically disadvantaged inner-city population | Tests had a fast turnaround time, simplicity, and mobility. Cost appeared to be a secondary consideration to end users. Most participants preferred low-cost devices | Of the 30 technicians enrolled, 26 (86.7%) had previous experience with a POC test, and POC tests were strongly preferred (83.3%) over standard laboratory-based NAAT (16.7%). Turnaround time (70.0%), sensitivity (36.7%), and ease of use (23.3%) were the parameters perceived to be of highest importance in an STI diagnostic test. Chlamydia (96.7%) and gonorrhea (96.7%) were identified as the most helpful targets for POC tests. | [25] |
| Wyatt et al. (2020) | Patients' experiences and attitudes of using a secure mobile phone app for medical photography: Qualitative survey study | To understand the perceptions, attitudes, and experiences of patients who were photographed using a mobile point-of-care clinical image capture app. | USA | Survey | Mobile point-of-care clinical image capture app | Mayo Clinic Minnesota<br>300 adult patients or parents of pediatric patients | 16% (10/62) of patients gave photographs to obtain advice from a specialist. 74% (51/69) would recommend medical photography to others and 67% (46/69) thought photos favorably affected their care. Patients were indifferent about the type of device used (mobile device vs professional camera; 40/69, 58%) or the identity of the photographer (provider vs professional photographer; 52/69, 75%). 90% (64/71) of patients found reuse of<br>photographs for one-on-one learner education to be acceptable.<br>Only 42% (30/71) of patients deeming reuse on social media for medical education as appropriate.<br>Only 3% [31] of patients expressed privacy or confidentiality concerns.<br>52% (33/63) of patients preferred to provide consent verbally, and 21% (13/63) did not think a specific consent process was necessary. | Patients were largely satisfied with clinical photography using the app. Some patients had the desire to incorporate personally taken photographs into their medical record and revealed evidence that some health care providers had resorted to using Photo Exam to take a photograph of a photograph displayed on a patient's smartphone screen as a workaround to incorporate these photographs. Many patients felt that verbal consent was adequate, verbal consent processes may be problematic in that they may not elicit specific permission for all potential reuses (i.e., medical education). | [31] |

The included studies were conducted in various countries (**Fig 2**). Three studies were conducted in the United States of America (USA) [25, 27, 31], one in Ghana [26], one in South Africa [24], one in the United Kingdom (UK) [29], one in Australia [32], and one in Germany [30]. A cohort study by Jacobson et al. (2020) also reported findings from the USA, Australia, Germany, Italy, Spain, Ireland, Brazil, Portugal, India, and Argentina [28].

**Fig. 2:**
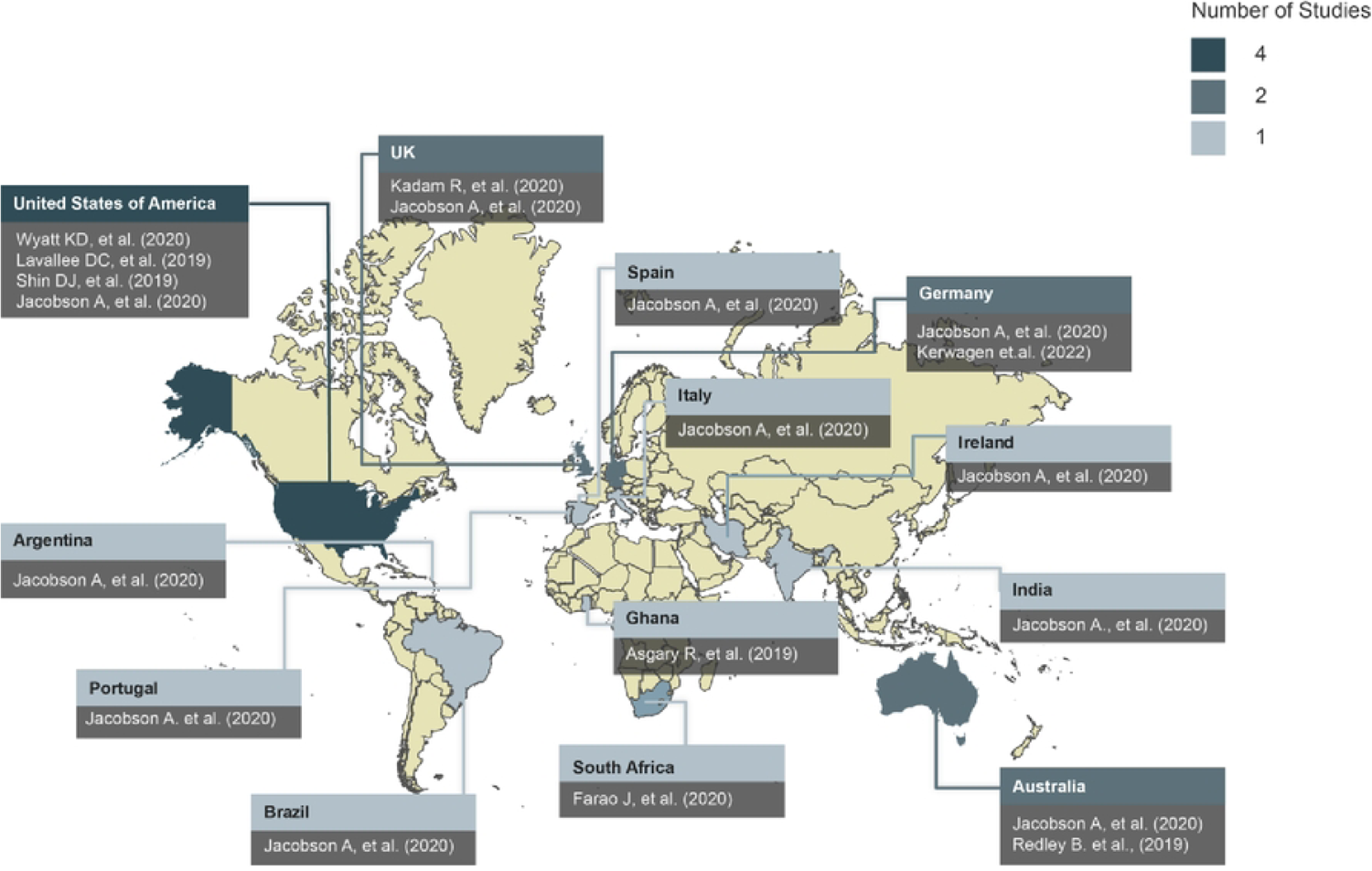
World map showing global evidence on integration of mobile-linked POC diagnostics in community-based healthcare with a focus on user experience identified in the included studies. The legend indicates the number of studies conducted per country.

The m-linked POC diagnostic technology presented in the included studies was focused on the following diagnostics (**Fig.3**): cervicography [26], tuberculosis [24], tumor stage [28], infectious diseases surveillance [29], speech recognition for diagnostic purposes [30], surgical site surveillance [27], neurocognitive disorders [32], chlamydia [25], and clinical image capture [31].

**Fig. 3:**
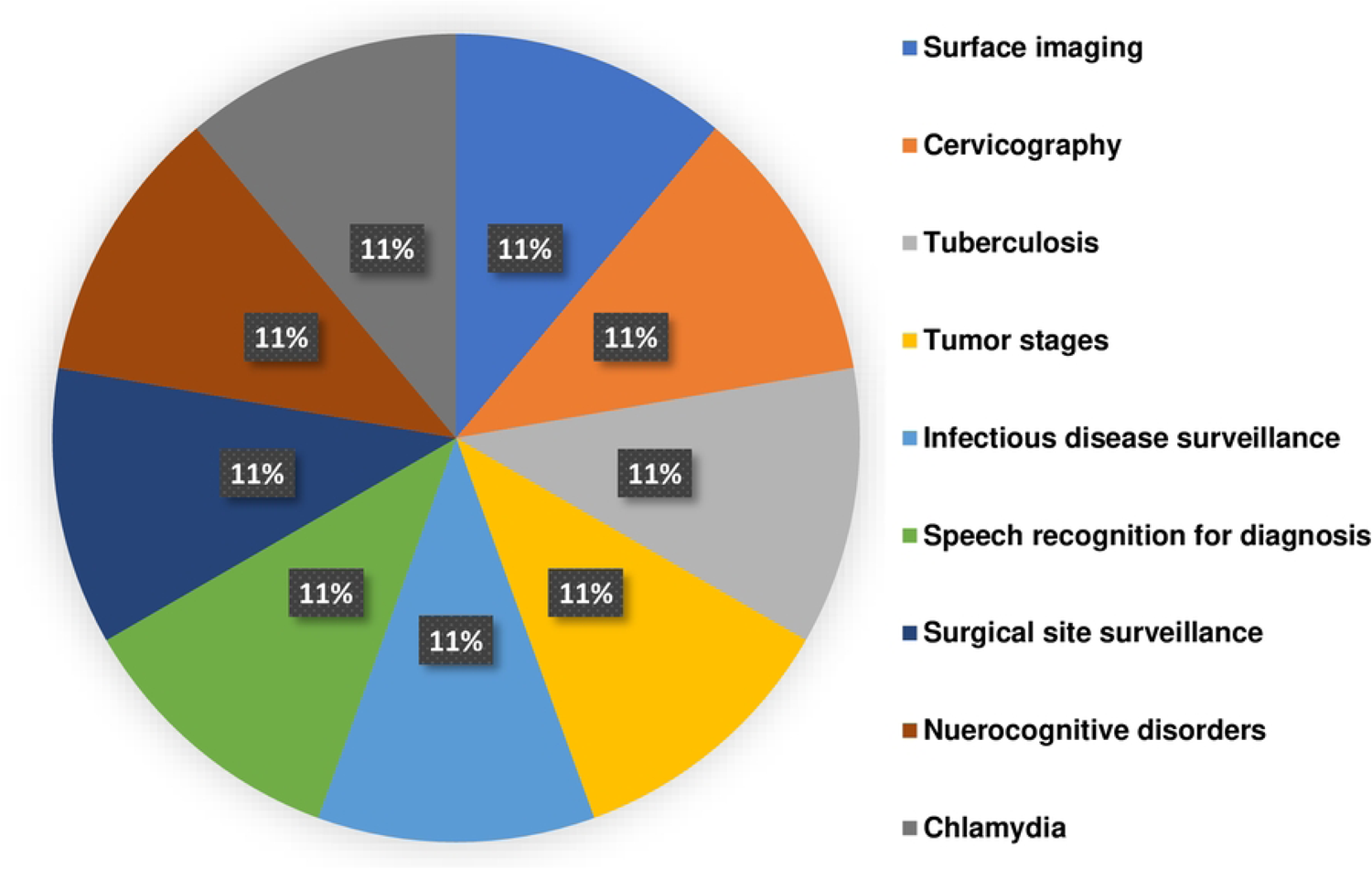
Target of m-linked POC diagnostic technology presented in included studies.

The included studies provided evidence of the integration of m-linked POC diagnostics in community healthcare (**Fig.4**). The community healthcare settings were emergency care units and departments [25, 31], community healthcare centers [26], developing and under-resourced contextual settings [24], the European Immuno-oncology Clinic CME community [28], infectious disease testing sites [29], a trauma and plastic surgery department [30], postoperative recovery [27], and hospital settings [32].

**Fig. 4:**
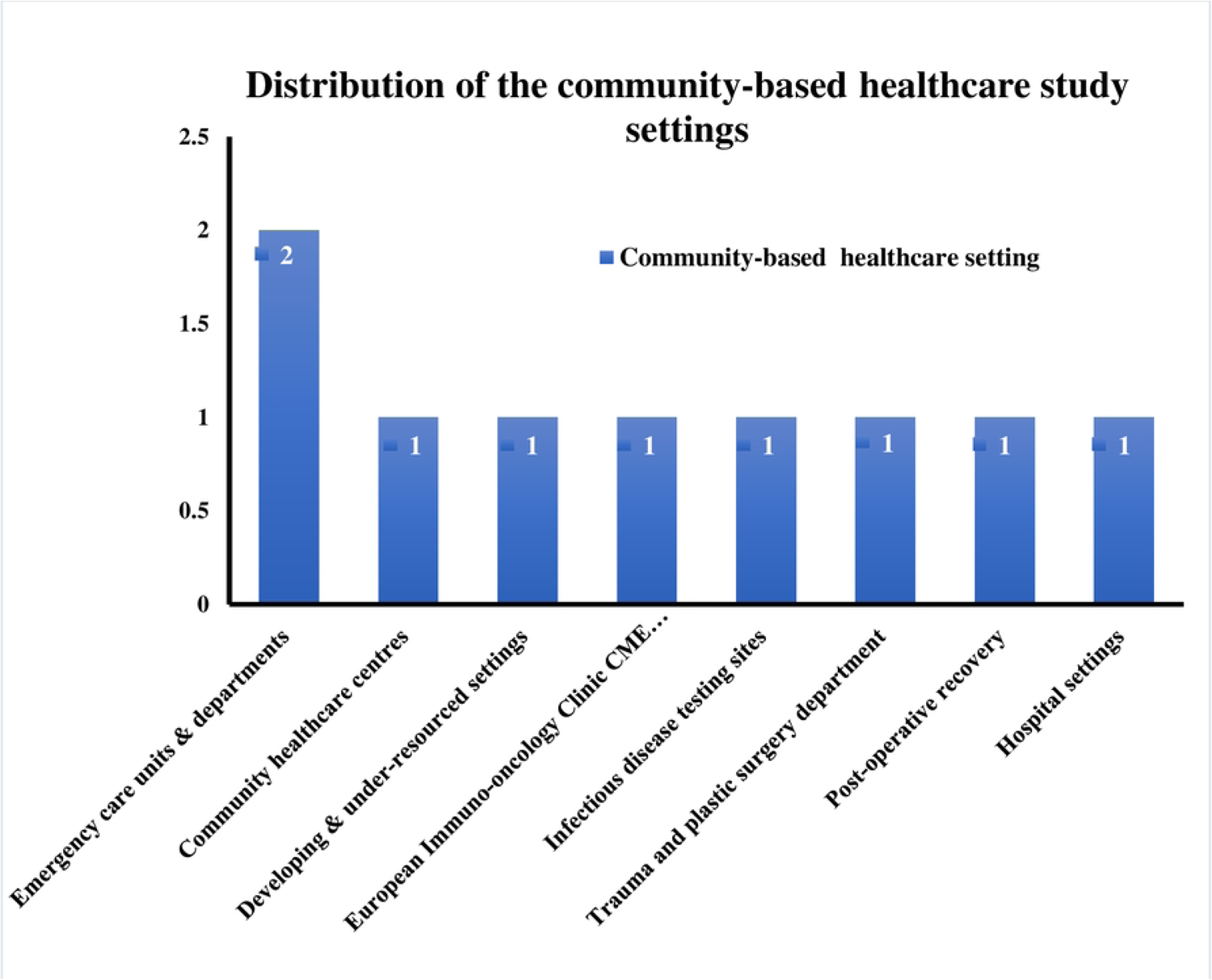
Distribution of community healthcare settings in which m-linked POC diagnostics were integrated.

The included studies focused on students (10%) [24], nurses (20%) [26, 32], patients (30%) [26, 31], healthcare professionals (20%) [28, 30], experts and key stakeholders (20%) [27, 29], as well as socioeconomically disadvantaged inner-city populations.

#### Quality of included studies

The quality of the nine included studies ranged from 72.5% to 100%. One study scored 72.5% (average quality), one study scored 95% (high quality), and the remaining seven studies scored 100% (high quality).

### Thematic findings

The following themes emerged from the included articles: approaches to m-linked POC diagnostic technology implementation, user engagement in community-based healthcare, and addressing limited user experiences in m-linked POC diagnostics.

### Approaches to implementing m-linked POC diagnostic technology

All nine studies showed evidence of steps taken to implement the developed technology [24–32]. In South Africa, Farao, Malila (24) explored a combination of user-centered approaches, specifically the “information systems research framework” and “design thinking,” to design a mHealth intervention for developing and under-resourced communities. They showed that user engagement was promoted by empathetic engagement with users, allowing for holistic and extensive communication [24]. Their findings were limited in that some end-users (healthcare workers) were not engaged throughout the study owing to constraints on their time and their availability [24]. This suggests that end-user engagement and experiences will depend on the state of the health system and the setting in which the intervention is piloted. The language used in the technology may also be a barrier to the use of m-link POC diagnostics suggesting a need for multi-lingual application [24]. Two studies conducted in the USA explored the perceptions, attitudes, and experiences of patients who were photographed using a mobile POC clinical image capture application [25, 31]. These studies concluded that end-user acceptability was linked to the level of involvement of end-users while developing the intervention. In Ghana, Asgary, Cole (26) investigated the acceptability and implementation challenges of smartphone-based training of community health nurses for cervical cancer screening in an urban setting. They noted that their findings could not be generalized to rural areas, which are markedly different from urban areas in terms of access to social and healthcare resources [26]. Jacobson, Macfarlane (28) tested the feasibility of integrating a mobile decision-support application into a multicomponent continuing medical education initiative to develop the competence of clinicians at POC in Australia, Portugal, Italy, Ireland, Argentina, Spain, Brazil, USA, India, Germany, and the UK, demonstrating that the strengths and limitations of such support tools need to be understood to advance the use of these resources in practice. In the UK, Kadam, White (29) aimed to create a target product profile for a mobile application that would read rapid diagnostic tests to improve and strengthen the surveillance of infectious diseases. They concluded the sustainability of such an app could be ensured by including additional languages, running several rapid tests simultaneously, affordability and multiple mobile device compatibility, reliable personal data security, and obtaining input from participants.

### User engagement in community-based healthcare settings

Eight studies presented evidence of user engagement [24-29, 31, 32]. In Ghana, Asgary, Cole (26) engaged with nurses who reported that they learned more when working with real patients than when attending theoretical training. In Australia, Redley, Richardson (32) qualitatively explored the acceptability, usability, and feasibility of a mobile application to support nurses in their care for patients with neurocognitive disorders in hospital settings. They found that feasibility and usability were enhanced by the ease of navigation and clarity and utility of content, but the use of the intervention was hindered by unclear expectations, unfamiliarity, and device-related factors [32]. Nurse expressed that acceptability was enhanced by familiarity and perceived benefits but hindered by perceived increases in workload, inconsistent use, pressure to use the application, and resistance to change [32]. In the USA, Lavallee, Lee (27) reported feedback from medical personnel who strongly suggested that patient experience should be included in the co-design of mobile health tools for surgical site infection surveillance. Shin, Lewis (25) evaluated the end-user acceptability of an experimental POC test platform and determined that end-users preferred POC testing over laboratory testing, provided that the devices were affordable. According to end-users, the acceptability of POC diagnostics is influenced by remote testing, security, and privacy [29, 31].

Three studies reported poor usability and user engagement [24, 28, 29]. In the study by Farao, Malila (24), users were unsure about the meaning of the data captured using the m-linked POC diagnostic device and how to interpret it. Poor user experiences were attributed to the inability to follow instructions [24], language barriers [24] [29], as well as poor integration of medical records and providing limited information [28].

### Addressing limited user experience in m-linked POC diagnostics

Of the nine included articles, six provided evidence for addressing the limitations of the user experience of the m-linked POC diagnostic technology [24, 25, 27–29, 31]. Lavallee, Lee (27) demonstrated that engaging with end users plays a vital role when implementing new healthcare interventions. Actively engaging with patients and healthcare professionals in community-based healthcare setting ensures that patient experiences are acknowledged and incorporated [27, 29]. Developers should also consider user behavior such as frequency of use and what they would primarily use the technology for when determining optimal engagement [28]. Users should also be encouraged to provide feedback on their experience after engaging with the implemented technology [29]. Feedback from healthcare workers in the USA, revealed that the manner in which a new product is presented, including the availability of instructional materials, may affect end-user experiences [25]. End-users need to engage with the implemented technology easily and efficiently, thus implementation language, ease of following instructional materials, and simplicity of the technology are important considerations. Studies could also be designed to include a representative sample of participants across a range of settings to address limitations such as those reported by Farao, Malila (24).

## Discussion

We conducted a scoping review to systematically map evidence on the user experiences of mobile-linked POC diagnostics in community-based healthcare settings. Although there is much research on POC diagnostics [33–41] in this digital age, few studies have incorporated user experiences that would inform developers and ensure sustainable implementation of such diagnostic tools. This scoping review identified a lack of context-driven development and implementation [26], thus hampering the upscaling of the developed diagnostic tools.

Organizations such as the WHO have discussed upscaling of mHealth innovations for specific population groups [7], which would require contextual understanding of end-user experiences [8]. Our scoping review also revealed a gap in the usability of diagnostics within community-based healthcare settings [24]. Developers of m-linked POC diagnostics need to consider contextual factors such as language, connectivity, and availability of devices. If end-users cannot engage with the technology, they are unable to provide useful feedback which would be concerning for developers and implementers of the diagnostic tools. Engaging both patients and medical personnel is key to advancing mHealth in community-based healthcare.

We believe that approaches to user engagement should be clearly described to inform the ease of use of the technology. User experience in the context of mHealth technologies should always focus on meeting the needs of users [11], which relates to our findings [25, 31]. However, in some cases, POC diagnostic tools were developed in collaboration with experts and patients, but only health experts gave feedback to improve on technological development [34, 42]. Moreover, end users need to trust that their data will be secure, and this could only be achieved if they can engage with the technology at the development stages before it is implemented within their community healthcare setting [43].

### Strengths and limitations of the study

Our scoping review was not limited by language, publication, or study design. The search also included grey literature. The quality of all the included studies ranged from 72.5% to 100% which is considered average to high quality. We only found nine articles that met our inclusion criteria; for this reason, the scoping review may not be appropriate to inform the implementation of m-linked POC diagnostic technology. Further primary research is thus needed on user experiences of m-linked POC diagnostic technology in community-based healthcare settings.

### Implications for research

We only found nine articles reporting the user experiences of m-linked POC diagnostic technology, which were conducted in recent years. The studies reported the user experiences of both medical personnel and patients, which is important when developing m-linked POC diagnostic tools. Some studies described that the technology was not user-friendly as language and lack of instruction hindered the usability of the technology. The security of patient data was still in question. Future research should also focus on how to efficiently integrate m-linked POC diagnostic technology without affecting the workflow of nurses and doctors. POC diagnostic technologies should ideally improve the workflow of healthcare workers and not be disruptive, which could potentially result in improved acceptability and sustainable implementation. More research is needed in a variety of healthcare settings focusing on different disease diagnostics.

### Implications for practice

The studies included in this scoping review mainly focused on small scale communities which would hinder their upscaling to larger community-based healthcare environments. Ideally, the implementation of these tools should be expanded to include a larger group of end users and different clinics and hospitals in a specific geographical setting. Several studies suggested that there should be more engagement with patients and not only medical personnel. Patients need to trust that by using this technology, their healthcare and personal information will not be compromised in any way.

## Conclusions

The evidence mapped in this scoping review highlighted the need for more research into the user experiences of m-linked POC diagnostic technology in community-based healthcare settings. Rapidly advancing technology and the emergence of various diseases, necessitates the implementation of m-linked POC technology to efficiently diagnose and treat diseases in all healthcare settings. The sustainability and efficient implementation of such technology depends on how users experience the technology.

## Data Availability

All relevant data are within the manuscript and its Supporting Information files.

## Acknowledgments

The authors would like to extend their appreciation to the University of Pretoria Faculty of Health Sciences Library services for their assistance with optimizing the search strategy. The authors extend their appreciation to Dr. Cheryl Tosh for editing.

## Authors’ contributions

SRN and TM-T conceptualized and designed the study. SRN prepared the first draft of the study, and KG assisted with the pilot database search. KM, TD, and BM contributed to the included studies’ abstract, full article screening, and quality assessment. SRN and TM-T contributed to synthesizing data and designing the sifting and data extraction processes. ABT and TM-T reviewed the manuscript. All authors reviewed the draft versions of the manuscript and approved the final version of the manuscript.

## Availability of data and materials

The data reported and supporting this paper was sourced from the existing literature and is therefore available through the detailed reference list.

## Ethics approval and consent to participate

This paper is a scoping review that only relied on the review of existing literature. There were no animal or human participants involved in this study and the study is not conducted for degree purposes. Therefore, ethical approval is not required.

## Consent for publication

Not applicable

## Supporting information

**S1 File. Level of agreement calculation results.docx**

**S1 Table. The search Summary Table (SST) of the database search used for this study.docx**

**S2 Table. Full article screening results from Reviewer 1 and Reviewer 2.xlsx S3 Table. Methodological quality appraisal of included studies.xlsx**

**S4 Table. The excluded studies and reasons for exclusion.docx S5 Table. PRISMA-ScR checklist.docx**

